# Reverse transcription loop-mediated isothermal amplification combined with nanoparticles-based biosensor for diagnosis of COVID-19

**DOI:** 10.1101/2020.03.17.20037796

**Authors:** Xiong Zhu, Xiaoxia Wang, Limei Han, Ting Chen, Licheng Wang, Huan Li, Sha Li, Lvfen He, Xiaoying Fu, Shaojin Chen, Mei Xing, Hai Chen, Yi Wang

## Abstract

Given the scale and rapid spread of severe acute respiratory syndrome coronavirus 2 (SARS-CoV-2, known as 2019-nCov) infection (COVID-19), the ongoing global SARS-CoV-2 outbreak has become a huge public health issue. Rapid and precise diagnostic methods are thus immediately needed for diagnosing COVID-19, providing timely treatment and facilitating infection control. A one-step reverse transcription loop-mediated isothermal amplification (RT-LAMP) coupled with nanoparticles-based biosensor (NBS) assay (RT-LAMP-NBS) was successfully established for rapidly and accurately diagnosing COVID-19. A simple equipment (such as heating block) was required for maintaining a constant temperature (63°C) for only 40 min. Using two designed LAMP primer sets, F1ab (opening reading frame 1a/b) and np (nucleoprotein) genes of SARS-CoV-2 were simultaneously amplified and detected in a ‘one-step’ and ‘single-tube’ reaction, and the detection results were easily interpreted by NBS. The sensitivity of SARS-CoV-2 RT-LAMP-NBS was 12 copies (each of detection target) per reaction, and no cross-reactivity was generated from non-SARS-CoV-2 templates. Among clinically diagnosed COVID-19 patients, the analytical sensitivity of SARS-CoV-2 was 100% (33/33) in the oropharynx swab samples, and the assay’s specificity was also 100% (96/96) when analyzed the clinical samples collected from non-COVID-19 patients. The total diagnosis test from sample collection to result interpretation only takes approximately 1 h. In sum, the RT-LAMP-NBS is a promising tool for diagnosing the current SARS-CoV-2 infection in first line field, public health and clinical laboratories, especially for resource-challenged regions.

## INTRODUCTION

In late December 2019, an unexpected outbreak caused by severe acute respiratory syndrome coronavirus 2 (SARS-CoV-2, known as 2019-nCov) emerged in Wuhan, Hubei province, China, which had previously not been reported in animals or humans (*1*). By press time, the novel coronavirus (SARS-CoV-2) has resulted in a huge epidemic in China and other 88 countries/territories, and there have been 98,129 globally confirmed cases of SARS-CoV-2 infection (COVID-19), including 3, 380 deaths (World Health Organization, COVID-19 Situation Report-46) (*2*). As the current outbreak of COVID-19 continues to evolve, COVID-19 has become a serious global health concern because of the possible fatal progression and rapidly growing numbers of new cases (*3*). Herein, it is urgently necessary to devise as many specific detection assays as possible to provide early, rapid and reliable diagnosis of COVID-19.

Diagnosis of COVID-19 based on clinical symptoms, especially in the early stages of this disease, is extremely difficult as there are no characteristic initial manifestations of COVID-19 (*4*). Although genome sequencing had high accuracy for diagnose of COVID-19, it was not applicable in rapid diagnosis for clinical large-samples because of its longer sequencing time and high requirements for experimental equipment (*5*). In the current outbreak of COVID-19, real-time reverse transcription polymerase chain reaction (rRT-PCR) was employed to detect SARS-CoV-2 in public health and clinical laboratories because it a specific and sensitive diagnostic method for detection the novel coronavirus (*6*). However, rRT-PCR technique strongly relies on complex apparatus, skilled personnel and a stable power supply, the total run time of rRT-PCR test is several hours from the clinical specimens to result reporting. In particular, some regions where outbreaks of COVID-19 emerge usually do not provide sufficient infrastructure for good rRT-PCR diagnostic services, especially for field laboratories and resource-limited settings (*7*). Hence, there is an urgent requirement for easy-to-use, more rapid and simpler detection techniques for diagnosis of COVID-19.

Loop-mediated isothermal amplification (LAMP), which is the most popular isothermal amplification assay, is able to offer a diagnostic testing option for this scenario (*8*). The details of LAMP mechanism have been described by previous publications (*9*). Using the LAMP-based protocols, nucleic acid amplification is conducted under isothermal conditions (e.g., in a heating block) with high efficiency, specificity and speed, eliminating the use of precision thermal cycler for thermal change (*9, 10*). The LAMP technique is highly specific, because recognition of target sequence by six or eight independent regions is required (*11*). Thus far, LAMP combined with reverse transcription assay (RT-LAMP) had been developed for the detection of multiple respiratory RNA viruses (e.g., influenza viruses, middle east respiratory syndrome and severe acute respiratory syndrome coronavirus) (*12*). Regarding these traits of LAMP technique, development of a LAMP-based assay for diagnosis of COVID-19 can overcome the shortcomings posed by rRT-PCR methods, and facilitates rapid diagnosis and surveillance of COVID-19.

Up to now, two informal published LAMP assays have been developed for diagnosis of COVID-19, and preliminarily evaluated using clinical or stimulated respiratory samples (*13, 14*). Unfortunately, only a genetic sequence (Open reading frame 1a/b; F1ab) was amplified and detected in the two systems, an unreliable diagnosis result may be obtained when the two COVID-19 LAMP assays detected a sample with high homology sequence of SARS-CoV-2 (e.g., bat severe acute respiratory syndrome-like coronavirus, GenBank KY417152.1). Traditional monitoring techniques (e.g., agarose gel electrophoresis, SYBR dyes and PH indicator), which were non-specific for COVID-19 LAMP products, were employed for confirming the two COVID-19 LAMP results. In addition, the electrophoresis is a time-consuming and tedious procedure, and the judgment of color change of reaction vessel by unaided eye is potentially subjective. Therefore, there is a continuous command for devising the new LAMP-based assays, which are capable of simultaneously detecting multiple targets of SARS-CoV-2, providing more rapid and objective result, and facilitating simpler diagnosis.

Here, a ‘one-step’ and ‘one-tube’ RT-LAMP coupled with nanoparticles-based biosensor (NBS) assay (RT-LAMP-NBS) was developed for diagnosis of COVID-19 (**Figure 1** and **2**). Two target sequences, including F1ab and nucleoprotein gene (np), were simultaneously amplified in an isothermal reaction, and detected in a test step. We will expound the basic COVID-19 RT-LAMP principle, optimize the reaction parameters (e.g., amplification temperature), and demonstrate its feasibility.

**Figure 1.**
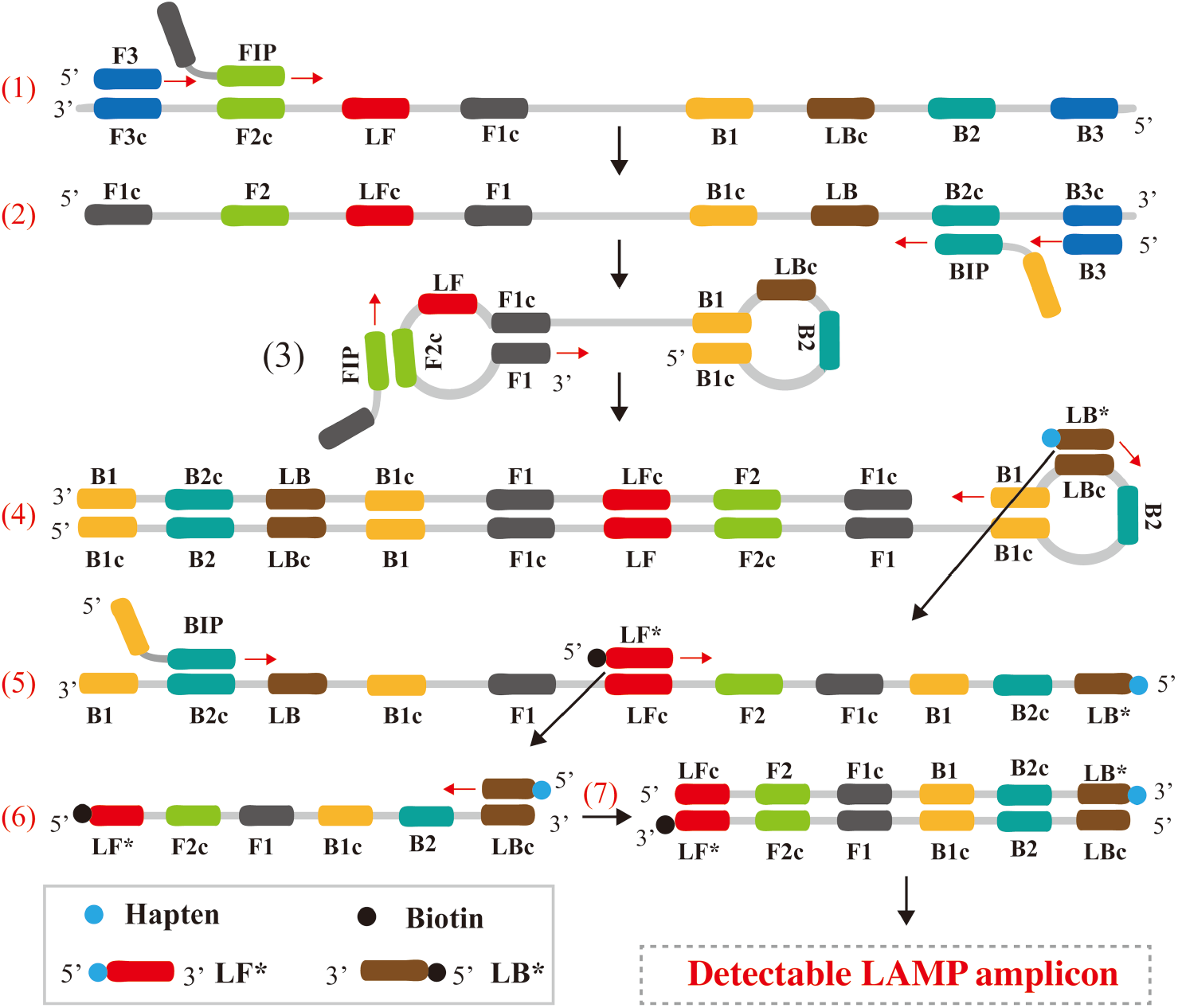
Outline of LAMP assay. **Top row**, outline of LAMP with LF* and LB*; **Bottom row**, schematic depiction of the new forward/backward loop primer (LF*/LB*). LF* was labeled with hapten at the 5’ end, and LB* was labeled with biotin at the 5’ end.

## RESULTS

### COVID-19 RT-LAMP-NBS design

In the LAMP system (**Figure 1**), FIP (forward inner primer) initiates the isothermal amplification, and the new strand derived from FIP primer was displaced by the F3 (forward primer) synthesis (**Step 1**). Then, 2 primers, including BIP (backward inner prime) and B3 (backward primer), annealed to the newly produced strand (**Step 2**), and displacement enzyme (*Bst* 2.0) extended in tandem generating a dumb-bell form product (**Step 3**). Thus, the stem-loop stem product can server as the template for the second stage of the LAMP reaction (exponential amplification). The LB* primer (backward loop primer), which was labeled with biotin at the 5’ end, could anneal to a distinct product derived from the exponential LAMP reaction stage (**Step 4**). The LB* product also severed as the template for next amplification by LF* (forward loop primer), which was modified at the 5’ end with hapten (**Step 5**). As a result, a double-labeled detectable product (LF*/LB* product) was formed, and one end of the LF*/LB* product was labeled with hapten, and the other end with biotin (**Step 6, 7**). One hapten is assigned to one primer set, which provide the possibility for multiplex LAMP detection.

A representative schematic of COVID-19 RT-LAMP-NBS assay were displayed in **Figure 2**. In the COVID-19 RT-LAMP system, fluorescein (FITC) was assigned to F1ab primer set, and digoxigenin (Dig) for np primer set. Hence, F1ab-LF* and F1ab-LB* primers were labeled at the 5’ end with FITC and biotin, and np-LF* and np-LB* for Dig and biotin, respectively (**Figure 2A**). With the assistance of AMV (avian myeloblastosis virus reverse transcriptase), the RNA (SARS-CoV-2 template) was converted to cDNA, which acted as the material for subsequent LAMP amplification (**Figure 2B**). After 40 min at 63°C, F1ab-LAMP products were simultaneously labeled with FITC and biotin, and np-LAMP products for Dig and biotin (**Figure 2C**).

**Figure 2.**
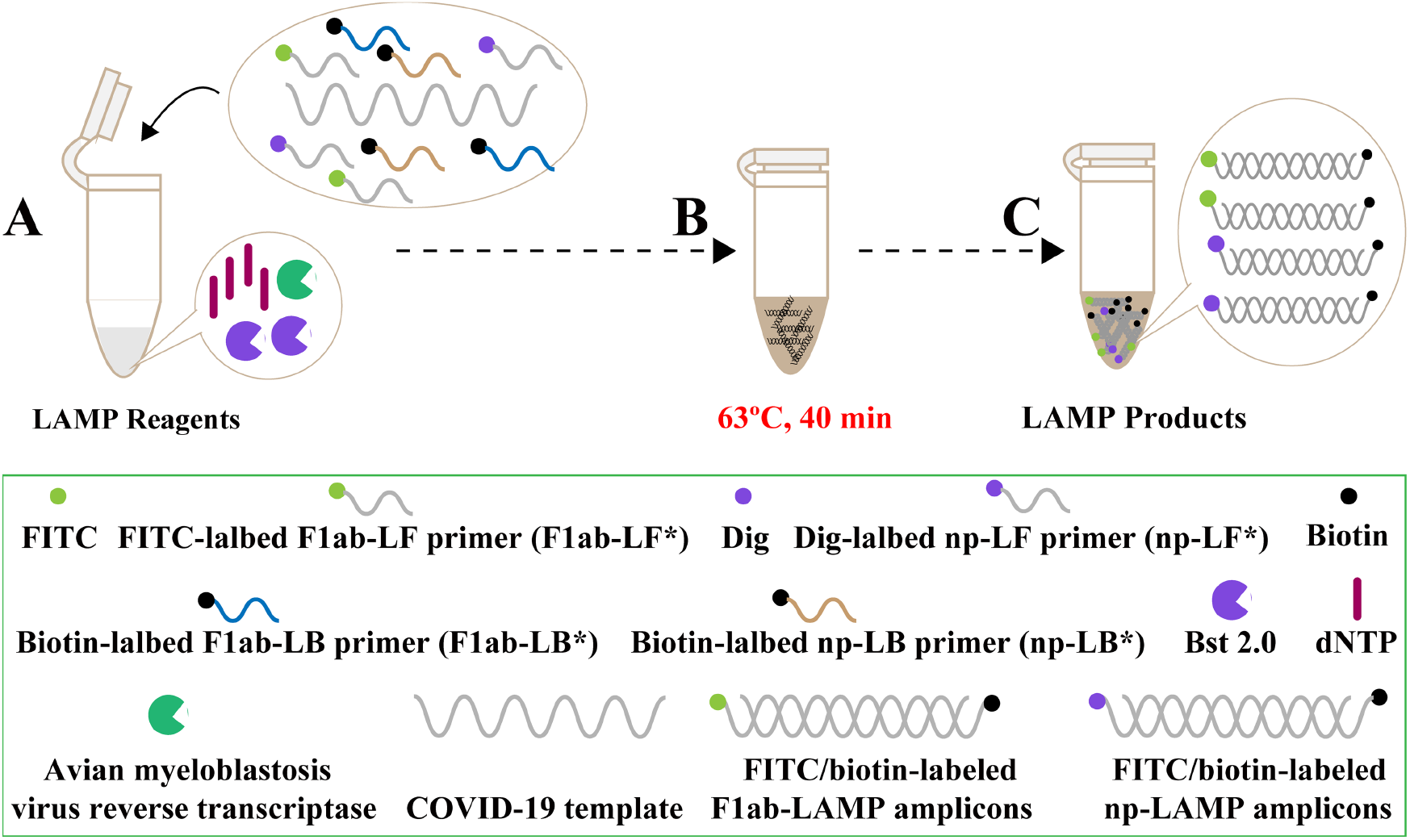
Mechanistic description of the COVID-19 RT-LAMP-NBS assay. **(A)**, Preparing the amplification mixtures. **(B)**, RT-LAMP reaction. **(C)**, The detectable COVID-19 RT-LAMP products were formed. F1ab-RT-LAMP products were simultaneously labeled with FITC and biotin, and np-RT-LAMP labeled with Dig and biotin.

### The principle of NBS visualization of COVID-19 RT-LAMP results

As shown in the **Figure 3**, the result readout of COVID-19 RT-LAMP assay was exhibited using the NBS. The details of NBS was shown **Figure 3A** (Seen in ‘Materials and Method’ section). For visualization of COVID-19 RT-LAMP results using NBS, aliquots (0.5 µl) of reaction mixtures were deposited into the sample region (**Figure 3B**, step 1), and an aliquot (80 µl) running buffer then was deposited on the same region (**Figure 3B**, step 2). At the detection stage, running buffer moves along NBS through capillary action, and rehydrates the SA-DNPS immobilized in the conjugate pad. The end of F1ab-RT-LAMP products labeled with FITC was captured by the anti-FITC antibody located in TL1 region (Test line 1), and the end of np-RT-LAMP products with Dig was captured by anti-Dig antibody located in TL2 region (Test line 2). The other ends of F1ab- and np-RT-LAMP products, labeled with biotin, bind streptavidin-conjugated color nanoparticles for visualization (**Figure 3B**, step 3). The excess streptavidin-conjugated color nanoparticles were captured by biotinylated bovine serum albumin immobilized in CL (Control line), which demonstrated the working condition of NBS (**Figure 3B**, step 3). The interpretation of the COVID-19 RT-LAMP results using NBS was shown in **Figure 3C**.

**Figure 3.**
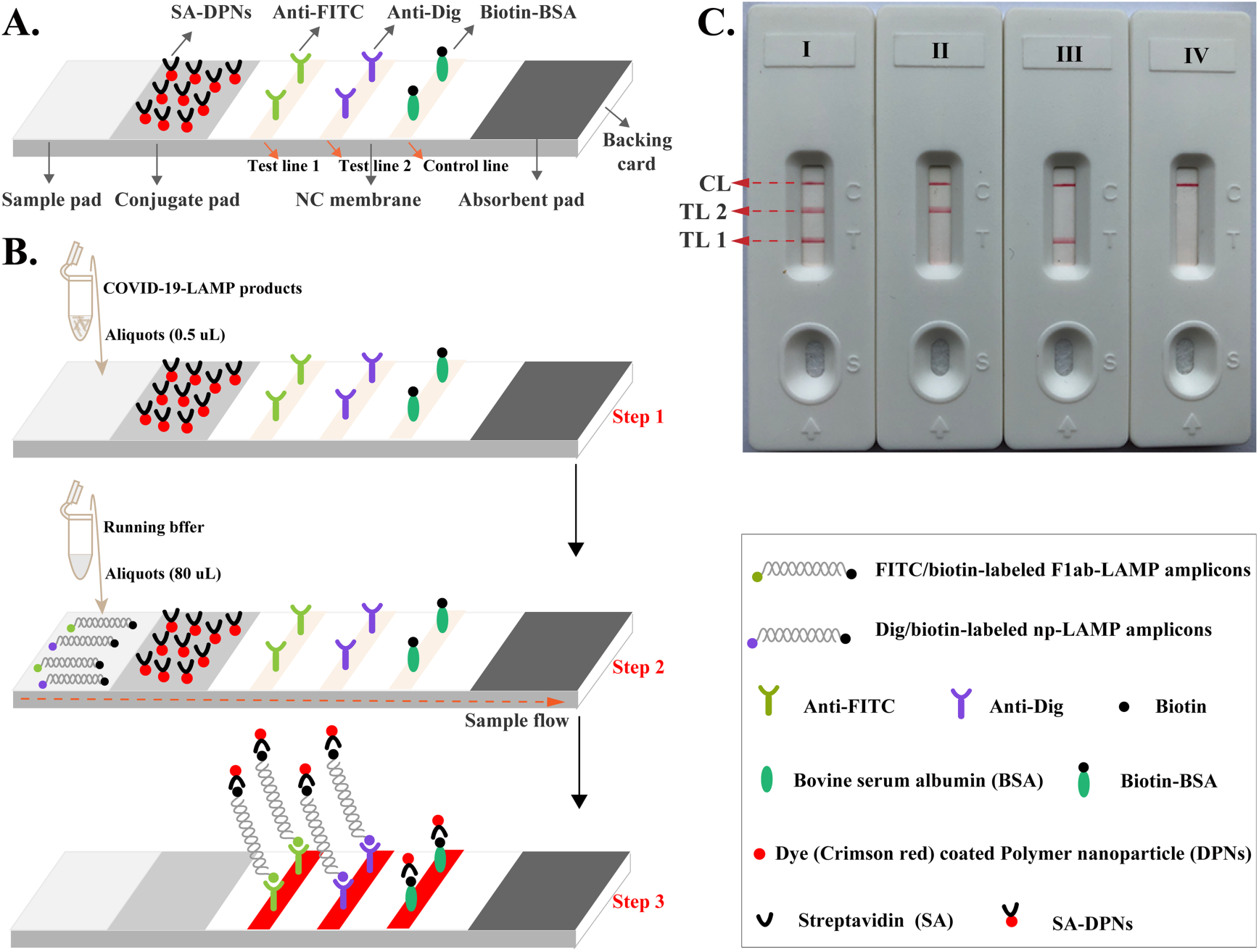
The principle of NBS for visualization of COVID-19 RT-LAMP products. (**A**), The details of NBS. (**B**), The principle of NBS for COVID-19 RT-LAMP products. (**C**), Interpretation of the COVID-19 RT-LAMP results. I, a positive result for F1ab and np (TL1, TL2 and CL appear on the NBS); II, a positive result for N (TL2 and CL appear on the detection region); III, a positive result for F1ab (TL1 and CL appear on the detection region); IV, negative (only the control line appears on the NBS).

### Confirmation and detection of F1ab-, np- and COVID-19 RT-LAMP products

The positive vessels of F1ab-, np- and COVID-19 RT-MCDA assay were visualized as light green using VDR (Visual detection reagent), while the reaction tubes of negative and blank controls remained colorlessness (**Figure S1**, top row). Using NBS, TL1 and CL were observed on the detection region for positive F1ab-RT-LAMP results, and TL2 and CL for positive np-RT-LAMP results. TL1, TL2 and CL simultaneously appeared on the detection region of NBS for positive COVID-19 RT-LAMP results. Only CL appeared on the analysis area of NBS for negative and blank controls of F1ab-, np- and COVID-19 RT-LAMP results (**Figure S1**, bottom row). These results indicated that F1ab- and np-LAMP primer sets were available for establish the COVID-19 RT-LAMP NBS assay for rapid and reliable detection of SARS-CoV-2. The parameter of optimal temperature for COVID-19 RT-LAMP technique also was tested, and reaction temperature of 63°C was used for performing the COVID-19 RT-LAMP amplification (**Figure S2** and **S3**).

### Sensitivity of COVID-19 RT-LAMP-NBS assay

The COVID-19 RT-LAMP-NBS was able to detect down 12 copies (each of F1ab-plasmid and np-plasmid) (**Figure 4**). Two target genes were detected and identified in a one-tube reaction (**Figure 4A**). The COVID-19 RT-LAMP results using NBS were in consistent with turbidity and VDR detection (**Figure 4B** and **4C**), while traditional monitoring techniques (VDR and turbidity) could not facilitate multiplex analysis. Furthermore, the sensitivity of COVID-19 RT-LAMP-NBS assay was in conformity with F1ab- and np-RT-LAMP assay (**Figure 4, S4** and **S5**).

**Figure 4.**
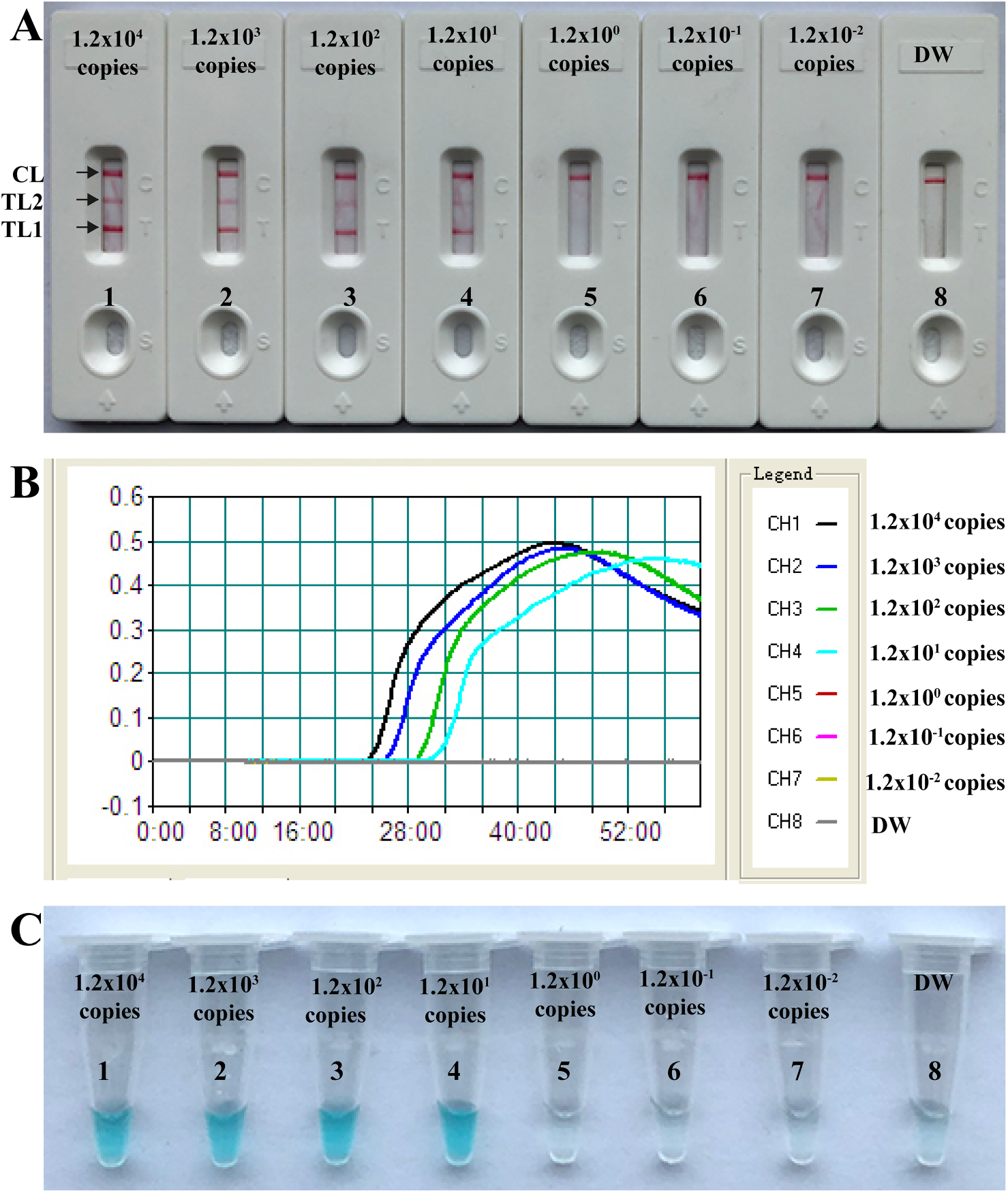
Sensitivity of COVID-19 RT-LAMP-NBS assay. **A**, NBS applied for reporting the results; **B**, Real-time turbidity applied for reporting the results; **C**, VDR applied for reporting the results. NBS (A)/Signals (B)/Tubes (C) 1-8 represented the plasmid levels (each of F1ab-plasmid and np-plasmid) of 1.2×10^4^, 1.2×10^3^, 1.2×10^2^, 1.2×10^1^, 1.2×10^0^, 1.2×10^−1^, 1.2×10^−2^ copies per reaction and blank control (DW). The plasmid levels of 1.2×10^4^ to 1.2×10^1^ copies per reaction produced the positive reactions.

The optimal duration time of COVID-19 RT-LAMP-NBS assay at the isothermal stage also was determined, and the template level at the detection limit appeared three red lines (TL1, TL2 and CL) when the RT-LAMP reaction was carried out only 30 min at 63°C (**Figure S6**). For the RNA template detection, a reverse transcription process (10 min) is essential, thus a COVID-19 RT-LAMP reaction time of 40 min was recommended for detection of clinical samples. Therefore, the whole diagnosis process of COVID-19 RT-LAMP-NBS, including sample collection (3 min), rapid RNA extraction (15 min), RT-LAMP reaction (40 min) and result interpretation (<2 min), was finished approximately 1 h (**Figure 5**).

**Figure 5.**
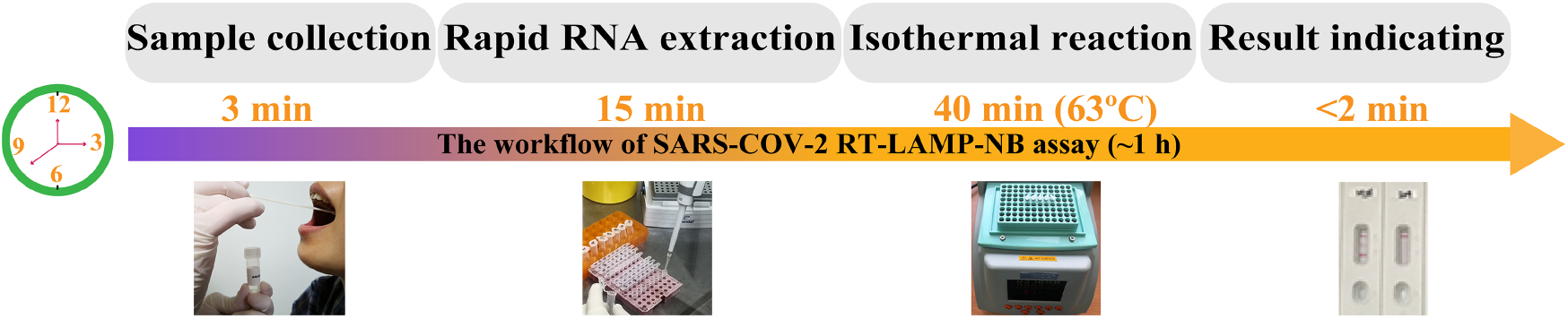
The workflow of COVID-19 RT-LAMP-BS assay. Four steps, including sample collection (3 min), rapid RNA extraction (15 min), RT-LAMP reaction (40 min) and result reporting (< 2 min), were required for conduct the COVID-19 RT-LAMP-NBS diagnosis test, and the whole process could be completed approximately 60 min.

### Specificity of RT-LAMP-NBS assay

The positive COVID-19 RT-LAMP-NBS results were obtained only from positive controls (120 copies each of F1ab-plasmid and np-plasmid) (**Table S1**). The negative results were observed in all pathogens of non-SARS-CoV-2 (virus, bacteria and fungi), in which only a red band (CL) was observed in the biosensor detection regions, indicating no cross-reaction with non-SARS-CoV-2 templates (**Table S2**).

### Application of the RT-LAMP-NBS assay in clinical samples

Of the total of 129 respiratory samples, which were initially analyzed using rRT-PCR in Sanya People’s Hospital in 2020, were enrolled in this report. Particularly, only the RNA templates were used after rRT-PCR performance. Among all the enrolled COVID-19 patients (33), the sensitivity of COVID-19 RT-LAMP-NBS assay 100% (33/33). The specificity was 100% (96/96) for COVID-19 RT-LAMP-NBS assay among non-COVID-19 patients, which were diagnosed as having pneumonia with confirmed pathogen in clinical laboratory of SanYa People’s Hospital. These preliminary results revealed that the proposed COVID-19 RT-LAMP-BS assay had a high sensitivity and specificity for diagnosis of SARS-CoV-2 infection.

## Discussion

An ongoing epidemic by SARS-CoV-2, starting in last December 2019 in Wuhan, China, is a huge public health concern (*15*). As of today (9 March 2020), 98 129 total confirmed cases have been documented, with 80 711 cases in China and the remaining cases distributed other 88 countries/regions in every continent (WHO, COVID-19 Situation Report-46) (*16*). Hence, there has been an immediate requirement for early, rapid and reliable diagnostic tests in the current outbreak. Such detection techniques are required not only in these countries where SARS-CoV-2 infection are spreading but also in countries/regions threated by SARS-CoV-2 infection, even in countries/regions where COVID-19 have not yet been emerged.

Here, we reported a novel LAMP-based test for simple, rapid and reliable diagnosis of COVID-19, name COVID-19 RT-LAMP-NBS. Our assay merged LAMP amplification, reverse transcription, multiplex analysis with nanoparticles-based biosensor, and facilitated the diagnosis of COVID-19 in a one-step, single-tube reaction. Only simple apparatus (e.g., a water bath or a heating block) were need to maintain a constant temperature (63°C) for 40 min. Compared with the developed COVID-19 RT-LAMP assays, the RT-LAMP results in this report were visually and objectively indicated by NBS, which was a simple and easy-to-use platform, avoiding the requirement of complex process (e.g., electrophoresis), special reagents (e.g., PH indicator) and expensive instrument (e.g., real-time turbidity) (*13, 14*). Total analysis procedure could be complete approximately 1 h, including sample collection (3 min), rapid RNA extraction (15 min), RT-LAMP reaction (40 min) and result interpretation (< 2 min). Considering these traits, COVID-19 RT-LAMP-NBS assay is a rapid, economical and technically simple method, offering a measure of practicality for field and clinical laboratories, especially for resource-challenged settings.

Two RT-LAMP primer sets, including F1ab-RT-LAMP and np-RT-LAMP primer sets, were specifically designed recognizing eight regions of target genes (**Figure 6**), guaranteeing the high specificity for SARS-CoV-2 detection. The data of analytical specificity revealed that no false-positive results were produced from non-SARS-CoV-2 templates, and positive results were obtained from positive control and SARS-CoV-2 templates (**Table S1**). Moreover, two targets (F1ab and np genes) could be simultaneously amplified and detected in a ‘one-step’ RT-LAMP reaction, which further guaranteed the assay’s reliability. Thus, our approach could effectively avoid the undesired results yielded from the developed COVID-19 RT-LAMP assays that only amplified and detected a target gene (e.g., F1ab) (*13, 14*).

**Figure 6.**
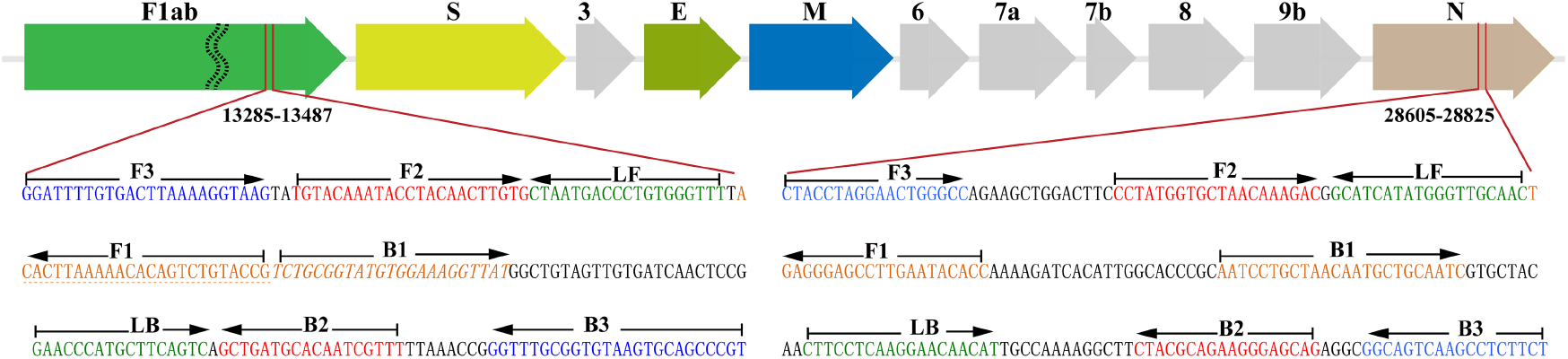
Primer design of COVID-19 RT-MCDA-BS assay. **Up row**, SARS-CoV-2 genome organization (GenBank: MN908947, Wuhan-Hu-1). The length of all genes was not displayed in scale. **Bottom row**, nucleotide sequence and location of F1ab and np gene used to design COVID-19 RT-LAMP primers. Part of nucleotide sequences of F1ab (**Left**) and N (**Right**) are listed. The sites of primer sequence were underline. Right arrows and Left arrows showed the sense and complementary sequence that are used. ^*^ Note: F1ab (Open reading frame 1a/b); S (Spike protein); E (Envelope protein); M (Membrane protein); N (Nucleoprotein); Accessory proteins (3, 6, 7a, 7b, and 9b).

The data of analytical sensitivity validated that RT-LAMP-NBS assay is sufficiently sensitive for diagnosis of COVID-19. Us this protocol, detection limit of COVID-19 RT-LAMP-NBS was 12 copies each of F1ab-plasmid and np-plasmid, which is in conformity with assay’s sensitivity generated from F1ab-RT-LAMP-NBS and N-RT-LAMP-NBS detection (**Figure 4, S4** and **S5**). The COVID-19 RT-LAMP-NBS assay did not improve or decrease the analytical sensitivity when compared with the signlex analysis (F1ab-RT-LAMP and np-RT-LAMP assays). In this report, we did not compare the sensitivity results obtained from COVID-19 RT-LAMP-NBS with rRT-PCR assay, because the quality of commercially available test kits for SARS-CoV-2 detection remains uneven. These commercial rRT-PCR assays used in Sanya People’s Hospital produce uninform results when they were applied to analyze the 10-fold diluted plasmid templates. For detection the RNA templates extracted from respiratory samples, 100% (33/33) of clinical samples examined by rRT-PCR exhibited completely consistent diagnosis, and analytical specificity was also 100% (96/96) for COVID-19 RT-LAMP-NBS when analyzing the RNA templates from non-SARS-CoV-2 infection patients.

## Conclusion

The one-step single-tube COVID-19 RT-LAMP-NBS assay devised in this report offers an attractive diagnosis tool for SARS-CoV-2 detection. The nearly equipment-free platform of COVID-19 RT-LAMP-NBS makes it applicable for resource-limited laboratories (e.g., field laboratories), and the diagnosis results are easy to interpret. The high specificity, sensitivity and feasibility of COVID-19 RT-LAMP-NBS assay for detection of SARS-CoV-2, and its low cost and ease of use make the target assay a valuable diagnosis tool for use in field, clinic, public heath and primary care laboratories, especially for resource-poor regions.

## Materials and Methods

### Construction of nanoparticles-based biosensor (NBS)

As shown in **Figure 3A**, NBS contains four components (a sample pad, a conjugate pad, a nitrocellulose membrane and an absorbent pad) (Jie-Yi Biotechnology). Rabbit anti-fluorescein antibody (anti-FITC, 0.2 mg/ml, Abcam. Co., Ltd.), sheep anti-digoxigenin antibody (Anti-Dig, 0.25 mg/mL, Abcam. Co., Ltd.) and biotinylated bovine serum albumin (biotin-BSA, 4 mg/mL, Abcam. Co., Ltd.) were immobilized at detection regions (nitrocellulose membrane, NC) as the test line 1 (TL1), test line 2 (TL2) and control line (CL), respectively, with each line separated by 5 mm. Dye streptavidin coated polymer nanoparticles (SA-DNPs, 129 nm, 10mg mL-1, 100mM borate, pH 8.5 with 0.1% BSA, 0.05% Tween 20 and 10mM EDTA) were immobilized at the conjugated regions. Thus, the NBS devised here can detect three targets (a chromatography control and two target amplicons). The assembled NBS were cut into 4-mm dipsticks, and dryly stored at the room temperature until use.

### Primer design

Two RT-LAMP primer sets (F1ab-RT-LAMP and np-RT-LAMP) were designed according the LAMP mechanism using a specialized software (PrimerExplore V5), which targeted F1ab and np gene of SARS-CoV-2 (GenBank MN908947, Wuhan-Hu-1) (**Figure 6**). Then, a Blast analysis of the GenBank nucleotide database was performed for the F1ab- and np-LAMP primers to validate sequence specificity. The more details of primer design, locations, sequences and modifications were shown in **Figure 6** and **Table S2**. All of the oligomers were synthesized and purified by RuiBo Biotech. Co., Ltd. (Beijing, China) at HPLC purification grade.

### Reverse transcription LAMP reaction (RT-LAMP)

The conventional RT-LAMP (F1ab- and np-RT-LAMP) was carried out in a one-step reaction in a 25-µl mixture containing 12.5 µl 2×isothermal reaction buffer [40 mM Tris-HCl (pH 8.8), 40 mM KCl, 16 mM MgSO_4_, 20 mM (NH4)_2_SO_4_, 2 M betaine and 0.2 % Tween-20], 8 U of Bst 2.0 DNA polymerase (New England Biolabs), 5 U of the avian myeloblastosis virus reverse transcriptase (Invitrogen), 1.4 mM dATP, 1.4 mM dCTP, 1.4 mM dGTP, 1.4 mM dTTP, 0.4 µM each of F3 and B3, 0.4 µM each of LF, LF*, LB and LB*, 1.6 µM each of FIP and BIP and template (1µl for the standard plasmid).

The COVID-19 RT-LAMP was also performed in a one-step reaction in a 25-µl mixture containing 12.5 µl 2×isothermal reaction buffer [40 mM Tris-HCl (pH 8.8), 40 mM KCl, 16 mM MgSO_4_, 20 mM (NH4)_2_SO_4_, 2 M betaine and 0.2 % Tween-20], 8 U of Bst 2.0 DNA polymerase (New England Biolabs), 5 U of the avian myeloblastosis virus reverse transcriptase (Invitrogen), 1.4 mM dATP, 1.4 mM dCTP,= 1.4 mM dGTP, 1.4 mM dTTP, 0.25 µM each of F1ab-F3 and F1ab-B3, 0.25 µM each of F1ab-LF, F1ab-LF*, F1ab-LB and F1ab-LB*, 1.0 µM each of F1ab-FIP and F1ab-BIP, 0.15 µM each of np-F3 and np-B3, 0.15 µM each of np-LF, np-LF*, np-LB and np-LB*, 0.6 µM each of np-FIP and np-BIP and template (1µl for the each standard plasmid, 5µl for samples).

The monitoring techniques, including real-time turbidity (LA-320C), visual detection reagents (VDR) and NBS, were employed for confirming the RT-LAMP reactions and optimizing the reaction parameters (e.g., reaction temperature and isothermal time).

### Sensitivity of the RT-LAMP-NBS assay

Two standard plasmids (F1ab-plasmid and np-plasmid) were commercially constructed by Tianyi-Huiyuan Biotech. Co., Ltd. (Beijing, China), which contain the F1ab and np sequences, respectively. Ten-fold serial dilutions (1.2×10^4^ to 1.2×10^−2^ copies) of F1ab-plasmid and np-plasmid were used to evaluate assay’s sensitivity. The plasmid concentration at detection limit level was employed for testing the duration time required by COVID-19 RT-LAMP-NBS assay.

### Specificity of the COVID-19 RT-LAMP-NBS assay

The specificity of the COVID-19 RT-LAMP-NBS assay was examined by detecting the templates extracted from various pathogens, including viruses, bacteria and fungi (**Table S1**).

### Feasibility of COVID-19 RT-LAMP-NBS using clinical samples

Respiratory samples were collected from COVID-19 infection patients in SanYa People’s Hospital, Hainan, which were defined according to standard diagnosis and treatment criteria of COVID-19 (Trial Version 6). The RNA templates extracted from respiratory samples were used after clinical and laboratory diagnosis, which was conducted using the RT-qPCR kit (Recommended by the China CDC). Collection of these RNA templates and analysis of them were approved by SanYa People’s Hospital. 5 µl RNA was used as templates for preforming the COVID-19 RT-LAMP-NBS test.

## Data Availability

all data of the study have been listed in this manuscript.

## Contributors

Yi Wang and Xiong Zhu conceived and designed this study. Xiong Zhu, Xiaoxia Wang, Licheng Wang, Limei Han, Huan Li, Ting Chen, Shaojin Chen, Mei Xing, Hai Chen and Yi Wang performed the experiments. Xiaoxia Wang, Xiong Zhu and Yi Wang analyze the data. Xiong Zhu, Xiaoxia Wang and Yi Wang contributed the reagents and analysis tools. Xiong Zhu, Xiaoxia Wang, Licheng Wang, Limei Han, Huan Li, Ting Chen, Shaojin Chen, Mei Xing and Hai Chen contributed the materials. Yi Wang conducted the software. Xiong Zhu and Xiaoxia Wang drafted the manuscript. Xiong Zhu and Yi Wang revised the manuscript.

## Ethical approval

This study was approved by the Ethics Committee of the Sanya People’s Hospital (SYPH-2019(41)-2020-03-06).

## Data sharing

No additional data available.

## Transparency declaration

The lead author and guarantor affirms that the manuscript is an honest, accurate, and transparent account of the study being reported; that no important aspects of the study have been omitted; and that any discrepancies from the study as planned and registered have been explained.

